# Development and Validation of a Highly Generalizable Deep Learning Pulmonary Embolism Detection Algorithm

**DOI:** 10.1101/2020.10.09.20210112

**Authors:** Ryan Schmid, Jacob Johnson, Jennifer Ngo, Christine Lamoureux, Brian Baker, Lawrence Ngo

## Abstract

Several algorithms have been developed for the detection of pulmonary embolism, though generalizability and bias remain potential weaknesses due to small sample size and sample homogeneity. We developed and validated a highly generalizable deep-learning algorithm, Emboleye, for the detection of PE by using a large and diverse dataset, which included 30,574 computed tomography (CT) exams sourced from over 2,000 hospital sites. On angiography exams, Emboleye demonstrates an AUROC of 0.79 with a specificity of 0.99 while maintaining a sensitivity of 0.37 and PPV of 0.77. On non-angiography CT exams, Emboleye demonstrates an AUROC of 0.77 with a specificity of 0.99 while maintaining a sensitivity of 0.18 and PPV of 0.35.

## Introduction

Venous thromboembolism (VTE) is a disease spectrum including both deep vein thrombosis and pulmonary embolism (PE). VTE is the third most common cause of acute cardiovascular disease following coronary artery disease and stroke (1). VTE is very common within the clinical realm with an incidence estimated to be between 1 and 2 cases per 1000 persons, ultimately projecting to approximately 300,000 to 600,000 cases in the United States annually (2–6). Pulmonary embolism is also a significant cause of morbidity and mortality with mortality rates historically reported as high as 15-30% (7–9). Over time, improved management of patients with PE has led to a reduction in mortality with rates now reported as low as 1.8% (10).

The prompt diagnosis and initiation of anticoagulation therapy have been shown to be associated with reduced mortality in patients with acute pulmonary embolism (11). Computed tomography pulmonary angiography (CTPA) has become the gold standard for the diagnosis of PE due to its high sensitivity and specificity, speed of image acquisition and wide availability (12–14). Thus, the radiologist plays a significant role in the diagnosis and timely treatment of patients with acute pulmonary embolism.

Today’s radiologist faces a number of challenges in providing rapid and accurate interpretations of imaging including massively increased imaging volumes (15–17). Increased work hours and study volumes per radiologist may increase error rates (18,19). They can also lead to increased turnaround times and negatively impact patient outcomes for urgent conditions like PE. There has been increasing effort to develop tools to address these demands (20). Interest in artificial intelligence (AI) and deep learning has proliferated in medicine and been shown to equal or exceed physician performance on several tasks (21–23). AI algorithms have been developed for a variety of specific tasks, including detecting intracranial hemorrhage on head CT exams (24), acute abdominal findings on CT exams (25), critical and urgent findings on chest radiographs (26)), and pulmonary embolism (27,28). However, the training and validation process has typically been limited to data sourced from a single or small group of hospitals. In fact, this has led to a disproportionate bias of the literature towards the use of data from a small group of states within the United States (29). In the current study, we aim to address some of these issues of generalizability and bias by developing an algorithm for the detection of pulmonary embolism using a large and diverse dataset sourced from over 2,000 hospital sites from all 50 states in the United States.

## Methods

All CT imaging and radiologic report data were obtained from vRad and were fully anonymized in a Health Insurance Portability and Accountability-compliant manner using proprietary software from vRad. This study was approved with a waiver of consent by an external Institutional Review Board, Western IRB. The vRad database was queried for all chest angiography protocols. For the purposes of this paper, no distinction was made between CTPA and other types of chest angiography. Because of the heterogeneity of contrast bolus timing, there was a fair amount of overlap between “late” CTPA, which mimics the timing of an aorta study and “early” aorta protocol studies, which mimics CTPA exams. A total of 18,372 examinations between March and April 2017 were used for training and validation of the deep learning algorithm. This was sub-selected from a larger set of data from the vRad practice to enrich the dataset with positive cases. From this set, 1,207 were labeled as positive and 17,165 were labeled as negative for PE. Bounding boxes were drawn around every instance of PE within the positive data set.

An additional 12,202 unique CT examinations, which cover the chest anatomy, including chest CTs and abdomen CTs, between July and September 2019 were collected for the test set. This was randomly sampled from a larger set of CT examinations that were collected within the vRad practice. The test set was 51.3% female with a mean age of 54.2. Of these, 7,462 cases were angiography cases while 4,740 cases were non-angiography cases. Data in this study were obtained across the United States from more than 2,000 hospital sites, representing nearly all brands and models of CT scanners. The majority of scans were obtained in the emergency department setting with the remainder obtained from inpatients and outpatients.

A proprietary ensemble algorithm, Emboleye, was developed for the classification of PE from CT angiography protocols. Emboleye takes a set of DICOM files as input and generates a patient-level classification of whether an exam is positive or negative for PE. As part of this ensemble, the images were resized to 800 x 800 pixels, and a vascular window and level were applied to the images (L: 100, W: 700). A slice-wise convolutional neural network was trained using the RetinaNet architecture (30). A random forest model was applied to the output of this architecture to generate patient-level classifications.

The report for each examination in the test set was manually reviewed by one of two radiologists, LN or JN, for whether the examination was positive or negative. They have 4 and 7 years of experience in radiology, respectively. At the time of report labeling, they were both blinded to the results of any of the relevant algorithms for any of these exams. Given differences in confidence conveyed by the interpreting radiologists in these reports, rules were established to determine how to perform such a binary classification. The following were labeled as positive exams: (1) clear diagnoses of pulmonary embolism (e.g., “Positive for pulmonary embolism in the right lower lobe.”), (2) chronic pulmonary embolism, (3) a focal finding is questioned without an alternative diagnosis being favored (e.g., “Questionable superior segment right lower lobe pulmonary embolism.”).

The following were labeled as negative exams: (1) clear diagnosis of no pulmonary embolism (e.g., “No pulmonary embolism.”), (2) general limitation of the study (e.g. “No evidence of pulmonary embolus to the segmental level” or “Nondiagnostic study for pulmonary embolism.”), (3) an alternative diagnosis for a focal finding seems to be higher in probability (e.g. “The left lower lobe pulmonary artery contrast bolus fades out in the left lung base. May be related to impeded flow in the atelectatic lung; however, pulmonary embolism cannot be excluded.”).

Finally, the performance of the Emboleye was evaluated. Evaluation metrics included model sensitivity, specificity, positive predictive value (PPV) and negative predictive value (NPV). The receiver operating characteristic (ROC) curve and area under the ROC curve (AUROC) were then determined. Misclassifications by Emboleye were subsequently evaluated to identify any underlying trends.

## Results

Within the test set of CT exams, the incidence of PE was determined to be 0.07 (515 positive and 6947 negative). The incidence of PE for non-angiography CT with contrast exams was 0.01 (65 positive and 4675 negative). Overall model performance for the detection of acute pulmonary embolism is shown in Table 1 and Figure 1. The model AUROC was 0.79 for chest angiography exams and 0.77 for non-angiography CT with contrast exams. To identify PE cases with high confidence, we used a high specificity threshold. On angiography exams, we achieved a specificity of 0.99 and positive predictive value of 0.77 while retaining a sensitivity of 0.37 and negative predictive value of 0.96. On non-angiography CT with contrast exams, we achieved a specificity of 0.99 and positive predictive value of 0.35 while retaining a sensitivity of 0.18 and a negative predictive value of 0.99. Examples of correctly and incorrectly identified PE are shown in Figure 2.

**Table 1.** Model performance on angiography exams and on non-angiography exams. Statistics are chosen on the point on the ROC curve that corresponds to a specificity ∼ 0.99.

|  | **Chest Angiography** | **Non-angiography CT with contrast** |
| --- | --- | --- |
| **AUROC** | 0.79 | 0.77 |
| **Sensitivity** | 0.37 | 0.18 |
| **Specificity** | 0.99 | 0.99 |
| **PPV** | 0.77 | 0.35 |
| **NPV** | 0.96 | 0.99 |

**Figure 1.**
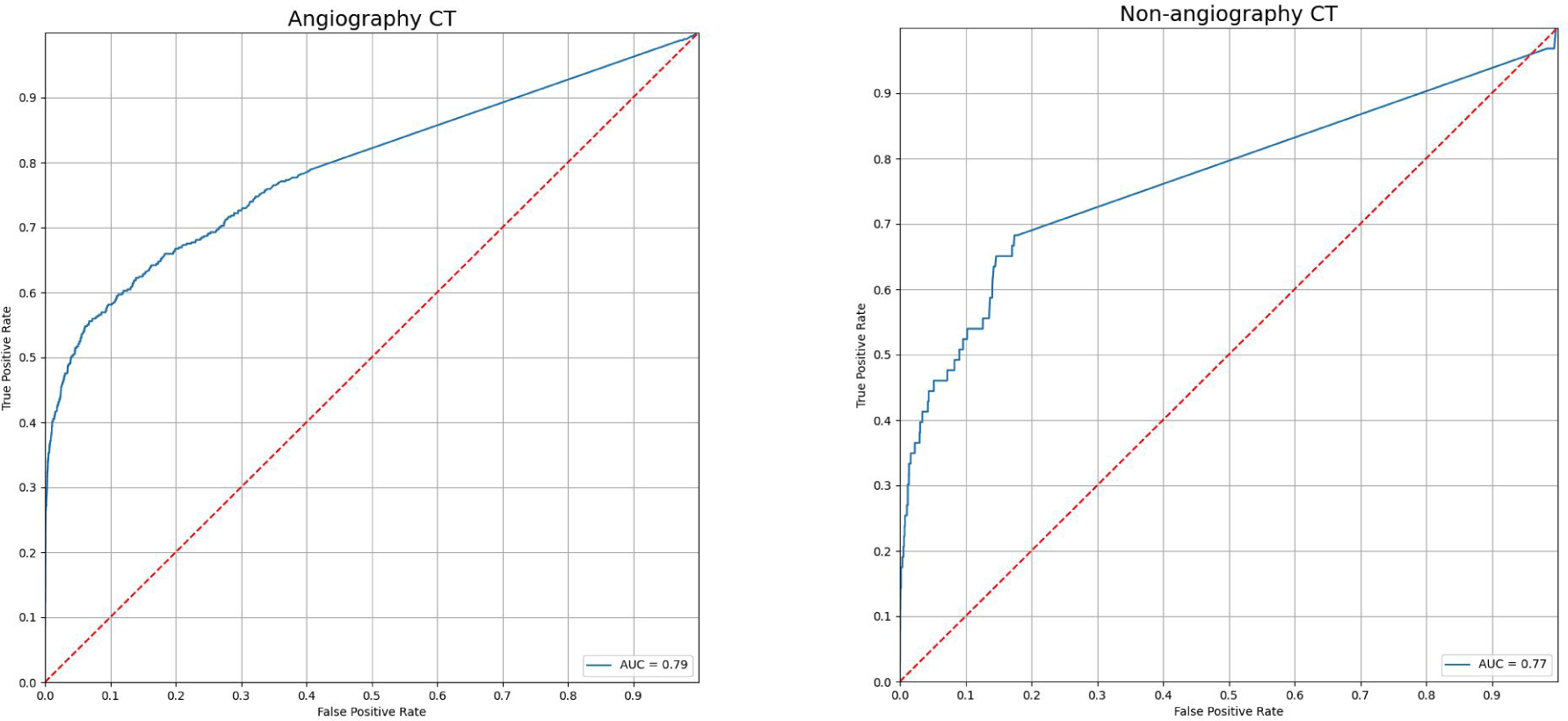
Receiver operating characteristic curves for angiography (AUC = 0.79) and non-angiography CT with contrast (AUC = 0.77).

**Figure 2.**
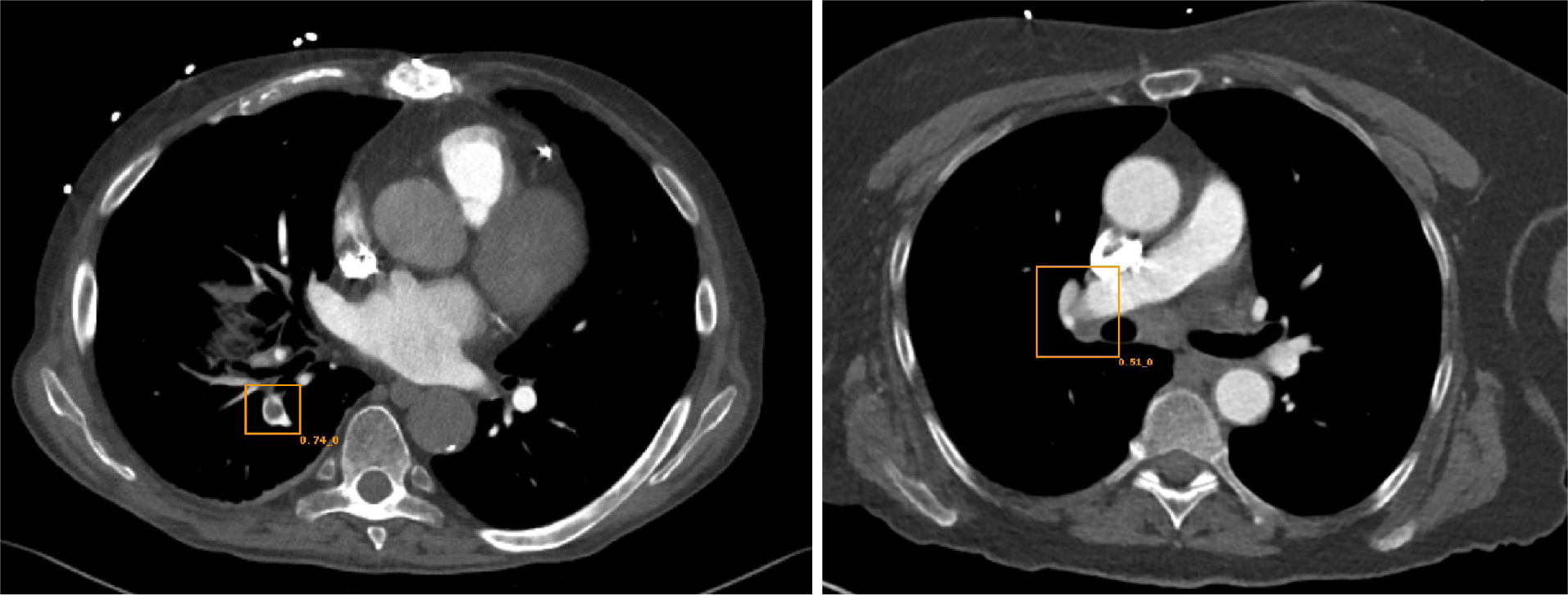
(a) True Positive. Emboleye correctly identifies segmental pulmonary embolism in the right lower lobe. (b) False Positive. Emboleye has a non-zero confidence rating regarding the absence of PE on this negative exam. The combination of a lymph node and streak artifact from the dense contrast in the adjacent SVC creates a confound for classification.

## Discussion

In this study, we have developed a deep learning-based model for the detection of pulmonary embolism on contrast-enhanced CT studies within cohorts of patients suspected and not suspected to have pulmonary embolism. To optimize the utility of Emboleye for clinical purposes, we established an extremely high decision confidence threshold, which sought to maximize specificity. To this aim, we believe that we have achieved start-of-art performance at the 0.99 specificity point compared to previous recent work on PE detection, which previously achieved a maximum sensitivity of 0.32 at this point (27,28). For a pathology with a relatively low incidence (0.07 on angiography exams and 0.01 on non-angiography), even a modestly high specificity would result in a large number of false positives. For example, in a cohort of 10,000 patients, an algorithm with a specificity of 0.80 would result in 1,860 and 1,980 false positives for angiography exams and non-angiography CT with contrast exams, respectively.

If the intended clinical use of such an algorithm were worklist prioritization, then such a system would be flooded with false positives, which would serve to deprioritize many other studies. This could be detrimental because some of these other studies could have critical findings. In fact, false positives may have a more detrimental effect than false negatives, because false negatives would merely represent the standard of care – they would not be reprioritized on the worklist. Therefore, despite tuning the algorithm to relatively low sensitivity, our algorithm is able to achieve a PPV of 0.77 on angiography exams and 0.35 on non-angiography CT with contrast exams, which are both much higher than the baseline incidences of 0.07 and 0.01, respectively.

Take an alternate approach to Quality Assurance (QA) as a clinical use case. In this use case, algorithms would both analyze the images and the radiologists’ reports to look for discrepancies. It would flag studies where, say, a PE is identified in the imaging, but where no diagnosis of PE is made in the report. Unless such an algorithm had perfect accuracy, there would be a need for radiologists to review these cases to determine whether such cases represented an algorithm error or an initial radiologist interpretation error. Having a high specificity which leads to a high positive predictive value would be critical in this case to minimize the number of studies that would have to be reviewed. A secondary review of a large number of cases with a low yield could be too costly for practices to implement on a regular basis.

From a statistical standpoint, these clinical considerations elicit an important point concerning the AUROC metric. Although such a metric is very often useful in measuring the effectiveness of different classifiers, it does not tell the entire story. For the clinical scenarios described above, the shape of the ROC curve may have a stronger bearing on the clinical usefulness of an algorithm. For example, even though the performance of Emboleye is similar across our two different datasets from an AUROC perspective (0.79 versus 0.77), the difference in performance when anchored at a specificity at 0.99 is more substantial. Therefore, it may be reasonable for algorithm-designers to not always seek to maximize AUROC, but may rather sacrifice AUROC in favor of achieving very specific points on a ROC curve that may have higher clinical yield.

In general, the development of an algorithm for the detection of PE is difficult given a variety of factors. Many anatomical structures can mimic the appearance of PE, including hilar lymph nodes, ribs, and contrast mixing in the SVC or pulmonary veins. Other factors may also contribute, including contrast-bolus timing, streak artifact, and motion artifact. Additional improvement of the algorithm will include the use of additional training data to robustly account for the full range of confounds as well as the use of more sophisticated 3D computation of the imaging datasets. Future work may also be directed towards the improvement of NLP models for the classification of PE based on text reports. Improvement of such algorithms can reduce the amount of human labor needed in monitoring the performance of these algorithms over time and for the identification of future training data sets.

Additional future work may also be aimed towards the review of the imaging findings for radiologist errors. In the current study, the diagnosis drawn from the report of the initial interpreting radiologist was taken as ground-truth. However, it is possible that radiologist over-calls and misses could influence the metrics of accuracy that we have reported in the current study. Though it would be infeasible in many circumstances to review the imaging findings from large datasets consisting of tens of thousands of exams, reviewing only discrepancies between imaging findings and report labels may be an effective strategy if an NLP algorithm is proven to have sufficient accuracy.

A general issue with deep learning algorithms has been their generalizability. A recent study demonstrated that although certain deep learning algorithms may have high performance for internal data sets, there is significant deterioration in performance for external data sets (31). Algorithms are typically trained using data from a limited source, and diversity in patient and scanner characteristics limits performance on disparate datasets. Emboleye was trained using data from over 2,000 hospital sites across the United States, covering the full gamut of patient populations and scanner types. A highly generalizable algorithm is particularly important for massively distributed practices like the one from which our data was obtained (vRad).

However, the current algorithm was trained solely from angiography examinations and then tested on both angiography and non-angiography CT with contrast exams. As expected, the accuracy of the algorithm decreases on the out-of-sample exam type. However, the performance degradation is not to such an extreme degree that would render such an algorithm useless. Given that the PPV of the algorithm on non-angiography CT with contrast is 0.35, the algorithm could still be an effective tool for clinical purposes. In fact, there may be mitigating circumstances, depending on the clinical scenario, which may result in improved clinical performance. For triage, there is a higher chance that such non-angiography studies would have a lower priority, so every correctly triaged exam of this type may have a larger benefit. Alternatively, for QA purposes, the rate of radiologist errors may actually be higher for incidental PE, which may ultimately lead to a clinically-significant yield. At any rate, future work will be aimed at including these non-angiography CT with contrast exams as part of training to even further increase generalizability.

## Data Availability

The data from this manuscript is proprietary and not publically available.

